# High-Sensitivity Pan-Cancer AI Assessment of Lymph Node Metastasis via Uncertainty Quantification

**DOI:** 10.64898/2025.12.23.25342895

**Authors:** Xiaodong Wang, Ying Chen, Xiaohong Liu, Cen Qiu, Hong Tang, Tinggui Huang, Siqi Guo, Sainan Ma, Mengjiao Cai, Qingyun Sun, Zichen Chang, Jinge Liu, Xiongjun Wang, Jinda Li, Wulei Qian, Biyu Wang, Boan Zhang, Chenguang Bai, Min Shi, Xinlei Zhang, Meng Li, Jiahai Wang, Bin Wang, Jinlu Ma, Lirong Ai, Shaoqing Yu, Liming Wang, Ninghan Feng, Xiyang Liu, Guanzhen Yu

**Author notes:** **Corresponding to**: Ninghan Feng, Xiyang Liu, Guanzhen Yu. These authors contributed equally: Xiaodong Wang, Ying Chen, Xiaohong Liu, Cen Qiu, Hong Tang.

## Abstract

The histological heterogeneity of primary tumours across the pan-cancer spectrum poses a formidable barrier to accurate lymph node metastasis assessment, often causing AI systems to make “overconfident errors” on rare variants that lead to missed diagnoses. To address this, we present UPATHLN, a unified diagnostic platform that synergizes a pathology foundation model-based encoder with a decoupled uncertainty estimation mechanism. We developed and validated the system using a large-scale multicentre dataset of 26,229 lymph nodes from 14 distinct primary origins. In internal validation, UPATHLN achieved an area under the curve (AUC) of 0.986. Crucially, the uncertainty module functioned as a decisive fail-safe: by flagging potential false-negative predictions for mandatory pathologist review, it intercepted all missed diagnoses, securing 100% conditional sensitivity across both the development and independent test cohorts—even for tumours from seven unseen primary origins. Concurrently, this mechanism reduced the review burden on negative lymph nodes by 73.2%. Ultimately, UPATHLN sets a new benchmark for safety-critical AI, demonstrating that explicitly modelling uncertainty is key to unlocking reliable, workload-efficient diagnostics at the pan-cancer scale.

## Introduction

Cancer remains a leading cause of death worldwide^1^, primarily driven by metastasis^2,3^. Lymph nodes (LNs) are critical anatomical checkpoints in cancer progression^4^, serving as initial metastatic sites and immune regulatory hubs orchestrating host-tumour interactions^5^. Accurate assessment of lymph node metastasis (LNM) status is therefore paramount for prognostic assessment and therapeutic decision-making across diverse cancer types^6^. However, current clinical management relies primarily on crude quantitative approaches—predominantly pN staging^6^—which fail to capture crucial biological features such as lymph node ratio (LNR), metastatic tumour burden, and diverse spatial growth patterns of metastatic cells within LNs. Recent evidence explicitly links elevated LNR to decreased survival rates and shortened recurrence-free intervals^7^, while highlighting how spatial distributions of metastatic cells can modulate the immune microenvironment, potentially facilitating immune escape^8,9^. Despite these insights, pathologists now face an unprecedented surge in LN specimens due to rising cancer incidence^10^, rendering such detailed, multi-dimensional analysis practically unfeasible in routine workflows.

Deep learning has rapidly advanced pathological diagnosis, with numerous artificial intelligence (AI) systems gaining regulatory approval. However, existing AI approaches for LNM detection^11–16^ typically follow cancer-specific paradigms. This fragmentation hinders widespread clinical adoption by necessitating multiple, disparate systems^17,18^, conflicting with the need for operational efficiency and unified diagnostic standards. Moreover, tumour heterogeneity^19,20^ manifests as a “long-tail distribution,” where common metastatic patterns coexist with diverse rare variants. Standard deep learning models often struggle with these outliers, prone to “overconfident errors” where the system makes incorrect predictions with high certainty on out-of-distribution data. The reliable detection of these rare but clinically significant variants is crucial for patient safety^21,22^, yet remains challenging even with large datasets. While emerging foundation models^23–28^ offer powerful representational capabilities, their effective application across the pan-cancer spectrum requires substantial annotation resources. Thus, there is an urgent need to transcend these limitations and establish a unified, robust, and safe diagnostic framework capable of not only high sensitivity but also reliable failure awareness all cancer types.

To address these challenges, we developed UPATHLN, a unified, uncertainty-aware AI platform integrating a pathology foundation model with uncertainty-guided active learning and robust uncertainty estimation. Trained and validated on an extensive multi-centre, multi-cancer LN dataset, this study demonstrates UPATHLN’s capacity for highly sensitive and safe pan-cancer LNM detection with robust generalization. Ultimately, this study represents a critical step towards establishing a unified, trustworthy diagnostic standard for AI-assisted pathology.

## Methods

### Dataset

Our dataset comprises retrospective multicancer LN pathology image data from three hospitals, annotated breast cancer LNs from the CAMELYON16 challenge^12^ (BLN16 dataset), and annotated gastric cancer LNs from previous work^13^ (GLN21 dataset). The dataset includes 2,738 patients (8,294 WSIs, 26,229 LNs) from 14 cancer types by tissue of origin (Fig. 1a). For the development cohort, we randomly selected 30% of WSIs from the stomach, breast, cervix, oesophagus, lung, colon, and ovary. Additionally, we included the BLN16 and GLN21 datasets in the development cohort. The remaining data were used as a multi-centre multi-cancer test cohort.

**Figure 1.**
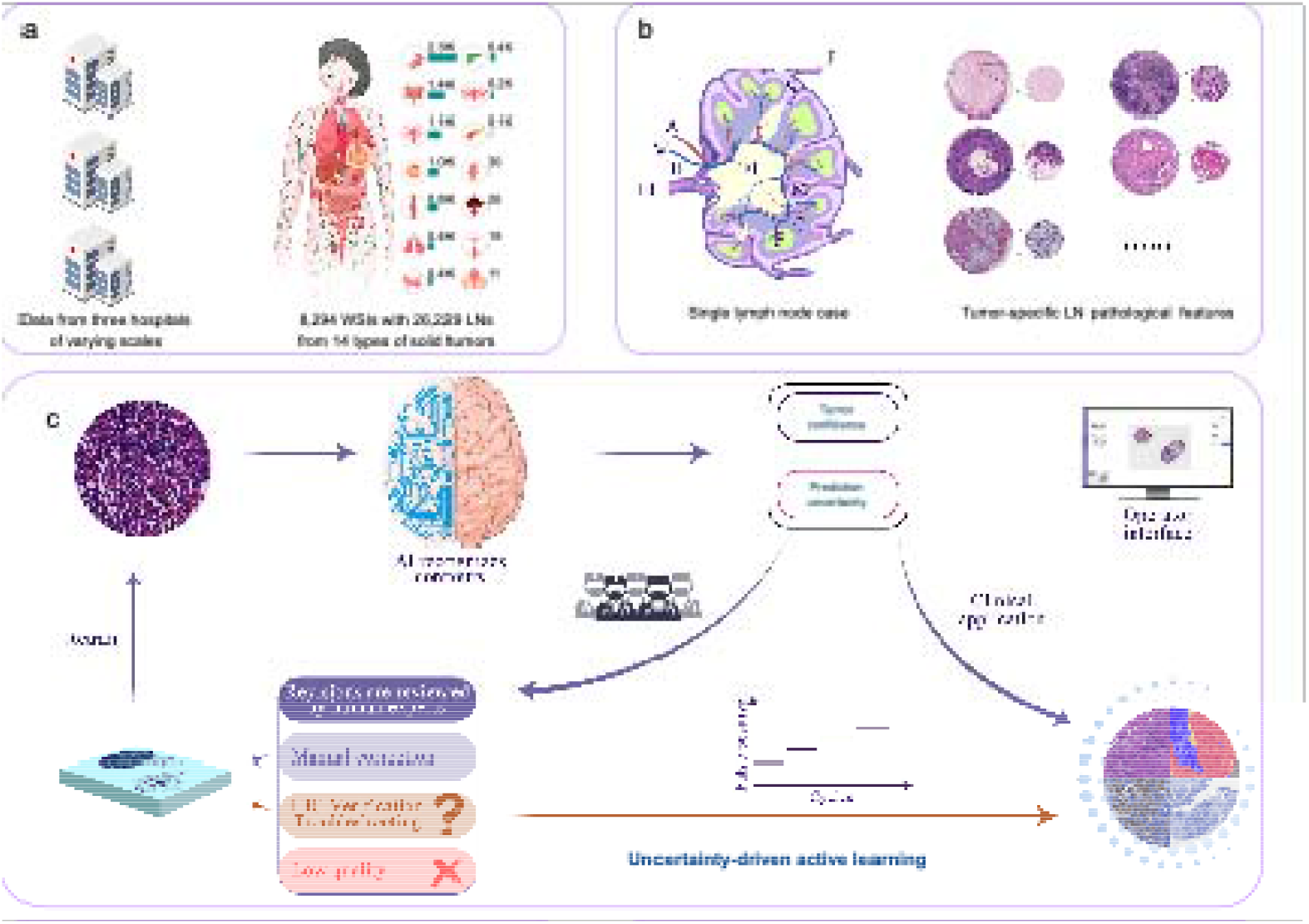
Development of clinically safe pan-cancer lymph node pathology diagnostic system. (a) Multi-center data composition: clinical samples from three hospitals of varying scales covering lymph nodes (LNs) from 14 primary sites. (b) Cancer-specific LN features: metastatic tumor cells in LNs often retain characteristics of primary tumors, with distinct histological patterns including thyroid cancer secretions, breast cancer lipomatosis, pancreatic cancer fibrosis, esophageal cancer keratin pearls, and colorectal cancer hemorrhage challenging pan-cancer analysis. (c) Uncertainty-guided active learning framework: system leverages clinical pathology database with iterative refinement where pathologists review high uncertainty (HU) predictions, verify challenging cases through IHC staining (CAM5.2, AE1/3), and exclude poor-quality samples. In clinical practice, uncertainty estimates guide pathologist review to prevent rare subtype misdiagnosis and enable continuous system improvement.

### Lymph Node Contour Recognition

To increase the training accuracy of LN metastasis detection models, we first developed an LN contour recognition model. Our model comprises two stages: initially, the YOLOv8-s model is employed for LN detection, and the detected regions are subsequently input into the nnUNet^29^ model for segmentation. We annotated 200 WSIs with 721 LNs for initial training. After initial training, we reviewed 918 WSIs (comprising 3,528 LNs) and reannotated 132 WSIs with incorrect predictions. We randomly selected 1,200 WSIs, 80% for training and 20% for testing, to retrain the YOLOv8-s model, which achieved a recall rate of 0.994 (95% CI: 0.990–0.998). We used the initial 200 WSIs along with the 132 fully corrected WSIs (comprising 1,245 LNs) for retraining the nnUNet model, which achieved a mean Dice score of 0.934 (95% CI: 0.923–0.945). The annotation and review process were conducted by two senior pathologists with over 15 years of experience: Y.C. and C.Q.

### Data Preprocessing for the Classification Model

Following the extraction of LN contour regions, we extracted image tiles of size 256×256 pixels at both 10x and 4× magnification from annotated regions, ensuring centre coordinates were aligned. Tiles were labelled positive if more than 25% of the area was annotated as tumour; otherwise, they were labelled negative. We employed a balanced sampling strategy to ensure an equal number of positive and negative tiles.

### Self-Supervised Pretrained Feature Extractor

We employed UNI^25^, a self-supervised foundation model for computational pathology (ViT-Large^30^ architecture), to extract powerful feature representations from HE-stained image tiles at both 10x and 4x magnification levels.

### Multiscale Feature Fusion and Classification Module

To capture both global tissue architecture and local cellular details, we developed a multiscale fusion module. Features extracted at 4x and 10x magnification were integrated using a cross-attention mechanism. This allows the model to learn the interplay between features at different resolutions. The fused features were then passed through self-attention layers and a final classifier to predict the probability of metastasis for each tile. During training, the parameters of the UNI pretrained feature extractor were fixed. We used AdamW for optimization with a weight decay of 0.05, a batch size of 64, and a cosine annealing learning rate schedule over 40 epochs.

### Uncertainty Estimation Module

We introduced an uncertainty estimation module to identify regions where the model’s prediction may be unreliable. This module was designed as an independent branch that takes the same internal features as the classifier. It was trained to predict the classification model’s error for a given region. This parallel design allows for efficient uncertainty quantification without compromising the primary diagnostic model’s performance or increasing inference time substantially. The module consists of three fully connected layers and was trained using an MSE loss function, with the AdamW optimizer over 10 epochs.

### Uncertainty-Driven Active Learning

We developed a human-in-the-loop active learning framework (Fig. 1c). We initialized the model using the GLN21 and BLN16 datasets. In subsequent iterations, unannotated WSIs from the development cohort with the highest model-predicted uncertainty were prioritized for review and annotation by senior pathologists (>15 years of experience, C.Q., J.L., C.B., M.S.). We performed four such iterations, reviewing a total of 600 WSIs. During the review process, pathologists performed immunohistochemical (IHC) staining on challenging cases, and cases that were CAM5.2 negative but suspicious were further validated via AE1/3.

### Multi-centre Clinical Validation

We conducted model validation using data from three centres. A dual-review process by pathologists (Y.C., C.Q., X.H.L.) was implemented to assess potential missed LNs and false positive/negative regions. Controversial WSIs underwent IHC verification (52 cases).

### Statistical Analysis

Detection and segmentation models used probability thresholds of 0.25 and 0.50, respectively. For metastasis classification, a threshold of 0.50 was applied. The uncertainty threshold was determined by balancing LN-level sensitivity and the number of HU regions. For performance evaluation, regions flagged as HU were considered positive for sensitivity calculations. Bootstrap analysis (1,000 iterations) was used to calculate performance metrics and their 95% CIs.

## Results

### UPATHLN Platform Development and Active Learning Strategy

We developed the UPATHLN platform using a human-in-the-loop active learning strategy (**Fig. 1**). We first collected diverse LN pathology data from three hospitals in China, representing varying levels of medical resources and patient populations. The dataset encompasses LNs from tumours originating in 14 organs (stomach, cervix, breast, thyroid, bile duct, lung, ovary, colon, oesophagus, pancreas, kidney, prostate, penis, and anus), totalling 2,738 patients contributing 8,294 haematoxylin and eosin (HE)-stained whole slide images (WSIs) with 26,229 LNs.

To ensure robust evaluation, we divided the data at the patient level into a development cohort and an independent test cohort. To comprehensively evaluate the model’s generalizability across cancer types, we strategically allocated certain cancer types exclusively to the test cohort, enabling rigorous assessment of performance on previously unseen cancer types.

We utilized a publicly available pathology foundation model, UNI^25^, as the feature extractor, upon which we built a classification model incorporating uncertainty estimation. During the training optimization phase, we initialized the classification model using previously annotated gastric cancer LNs (GLN21 dataset)^13^ and breast cancer LNs from the CAMELYON16 challenge (BLN16 dataset)^12^. Subsequently, we employed the uncertainty estimation module to evaluate predictions for unlabeled WSIs in the development cohort. WSIs ranked highest for high uncertainty (HU) regions were selected for pathologist correction and reintegrated into the dataset for model retraining. This iterative process was repeated five times until performance stabilized on the internal validation set.

### Uncertainty-Aware Classification Achieves High-Sensitivity LNM Detection

To ensure high-quality input, the extraction pipeline utilized YOLOv8-s for LN detection (Recall: 0.994; 95% CI: 0.990–0.998) and nnUNet for tissue segmentation (Dice: 0.934; 95% CI: 0.923–0.945), effectively excluding non-target tissues (Fig. 2a). The core classifier was subsequently refined through five rounds of uncertainty-driven active learning. Quantitative profiling of the prediction distribution confirmed that classification errors were predominantly sequestered within the HU tail, effectively isolating “hard examples” for targeted refinement. This optimization significantly enhanced discriminative power, improving the AUC from a baseline of 0.953 (95% CI: 0.932-0.974) to 0.986 (95% CI: 0.976-0.996), with consistent gains in both Positive and Negative Predictive Values (Fig. 2b-d).

**Figure 2.**
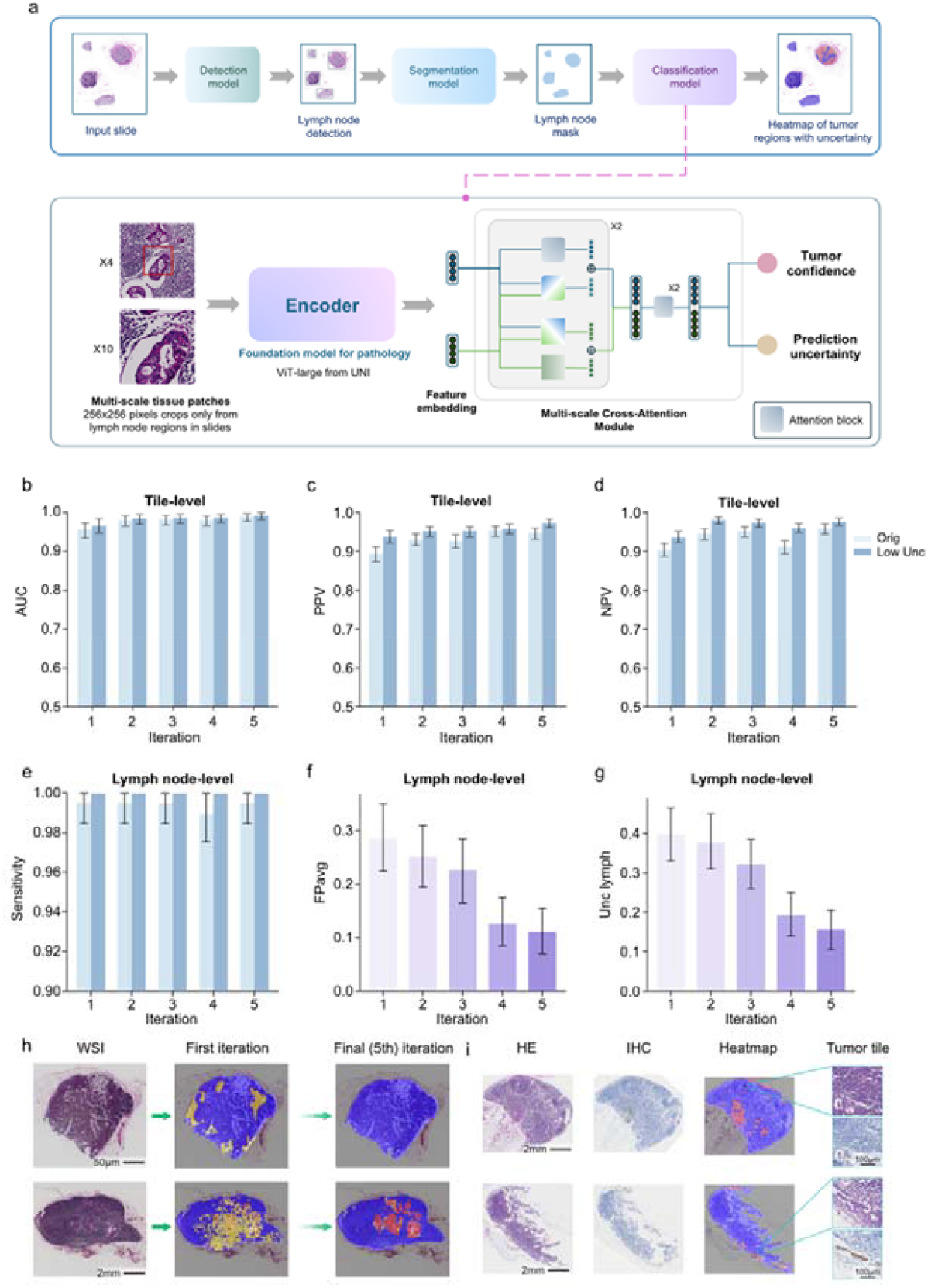
Deep learning framework for lymph node metastasis detection with iterative refinement. (a) System architecture overview: detection model identifies LN regions in WSIs; segmentation model delineates LN contours; classification model analyzes multi-scale tiles using UNI pathology foundation model, incorporating Cross-Attention for tumour confidence and uncertainty estimation. (b–e) Performance metrics comparing original and high uncertainty (HU)-filtered predictions across training iterations: tile-level AUC (b), PPV (c), NPV (d), and lymph node-level sensitivity (e). (f–g) Additional lymph node-level metrics: average false positives per slide (FPavg) and average high uncertainty regions per slide (HUavg). (h) Progressive improvement of prediction heatmaps across iterations. (i) Representative isolated LNM detection cases with immunohistochemistry (IHC) validation. In bar charts (b-g), 95% confidence intervals were derived from bootstrapping (1000 iterations). In heatmaps (h, i): yellow - HU regions, red - high tumour confidence regions, blue - LN regions.

To rigorously attribute this high performance to our architectural design rather than data accumulation alone, we benchmarked the model against alternative architectures trained on the identical dataset generated from the final iteration. Our multiscale architecture, leveraging the UNI foundation model, significantly outperformed both the single-scale UNI baseline (AUC=0.928, 95% CI: 0.910-0.946; *P*<0.0001) and the ImageNet-pretrained feature extractor (AUC=0.918, 95% CI: 0.901-0.935; *P*<0.0001). This distinct performance gap underscores the importance of incorporating multiscale context into foundation model-based workflows for robust metastasis detection.

Crucially, the uncertainty estimation served as a decisive fail-safe mechanism. While the optimized raw classifier achieved a robust intrinsic sensitivity of 0.996 (95% CI: 0.985–1.000), it failed to detect three instances of LNMs in its standalone configuration (false negatives) (Fig. 2e). However, the uncertainty module successfully captured the diagnostic ambiguity in these rare failure cases, flagging them for mandatory review. This capability to ‘rescue’ potential false negatives effectively neutralized the risk of missed diagnosis, underpinning the system’s 100% conditional sensitivity and ensuring that ambiguous but critical lesions were correctly routed for pathologist confirmation.

To facilitate clinical translation, we optimized the workflow to minimize review burden. Across iterations, the LN-level False Positive (FP) and HU rates steadily declined (Fig. 2f, g), reaching final values of 0.112 (95% CI: 0.070–0.155) and 0.156 (95% CI: 0.110–0.202) per slide, respectively. The model’s progressive refinement was visually corroborated by the prediction heatmaps (Fig. 2h). Furthermore, the system demonstrated precise identification of diverse metastatic burdens, including macro-metastases, micro-metastases, and isolated tumour cell clusters (Fig. 2i). Final validation by pathologists on the remaining development cohort confirmed the model’s robust performance, achieving a 100% positive detection rate with minimal review overhead (1% FP rate and 0.16% HU rate at the LN level).

### Validation of Pan-cancer Robustness and Safety

Performance evaluation on the independent test cohort confirmed the system’s robust discriminative power (Fig. 3). The classifier achieved a raw sensitivity of 98.7% and a specificity of 88.0% in its standalone configuration. While the raw model missed 60 LNMs, the uncertainty module functioned as a decisive fail-safe mechanism, successfully intercepting all 60 missed instances and flagging them as HU. This “rescue effect” ensured a 100% conditional sensitivity across the entire population, effectively neutralizing the risk of false negatives.

**Figure 3.**
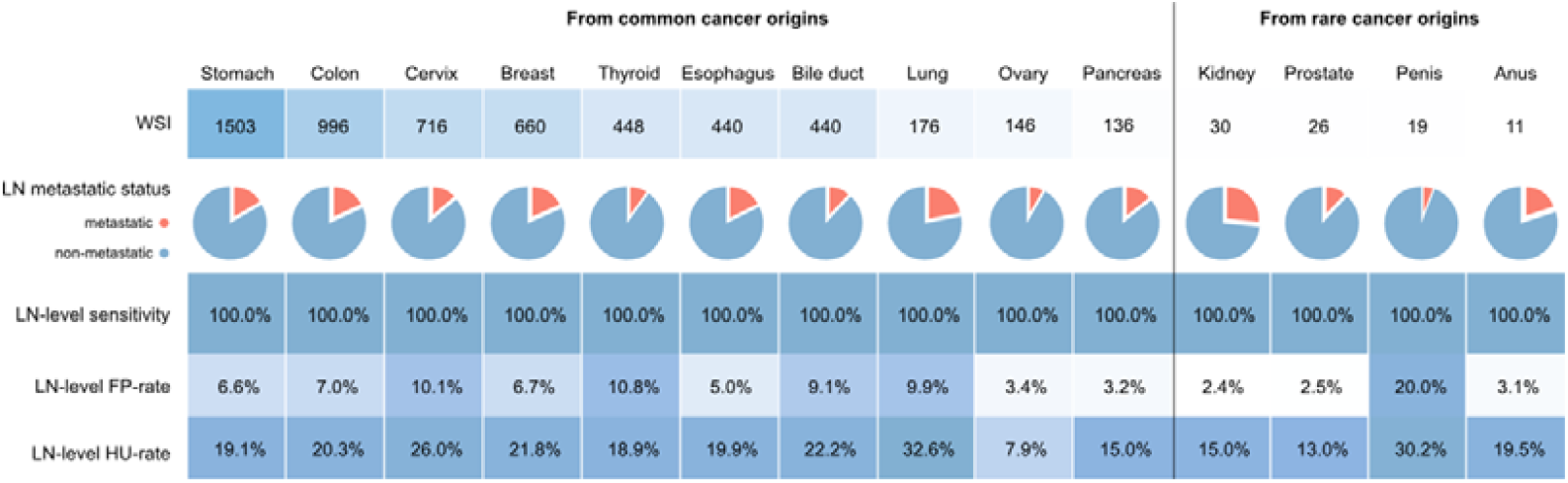
Performance evaluation of the classification model across multi-cancer lymph node datasets. In pie chart: blue - non-metastatic, red - metastatic.

The operational cost of this safety profile was quantified using the Review Burden Rate on Negative Cases (RBR-N). Accounting for the union of FP and HU predictions, the effective RBR-N was 26.8% (3,180/11,873). This indicates that 73.2% of negative LNs were safely excluded from manual review.

Subgroup analysis revealed remarkable consistency across organ origins, with a weighted average FP rate of 7.5% (SD: 2.1%) (Fig. 3). For the seven cancer types included in training, performance remained stable regardless of sample size. Notably, although Stomach cancer LNs were overrepresented in the training data (35.6%), their performance (HU rate: 5.2%; FP rate: 6.6%) closely mirrored the cohort average, indicating that the model maintained balanced discriminative power despite the non-uniform training distribution. While robust to sample size variations, the model exhibited sensitivity to specific non-neoplastic histological features. LNs from Lung and Cervix exhibited higher HU rates (32.6% and 26.0%, respectively). Pathological audit confirmed that in Lung cancer, these flags were predominantly triggered by anthracotic pigment, indicating that the model correctly identified non-malignant confounders as low-confidence regions.

Most notably, the system demonstrated exceptional adaptability to seven cancer types entirely unseen during training. For common unseen types, the model maintained low FP rates (e.g., Pancreas: 3.2%; Bile duct: 9.1%) without fine-tuning. Even for rare primary sites (kidney, prostate, penis, anus), the model achieved 100% conditional sensitivity. In challenging morphologies like Penile cancer, the model exhibited an adaptive increase in review rates (HU: 30.2%; FP: 20.0%), reflecting a cautious inference strategy that prioritizes safety in out-of-distribution domains.

### Interpretability and Pathological Characterization of Uncertainty

To elucidate the interpretability underlying this robust performance, we deconstructed the uncertainty landscape. Multiscale feature fusion dimensionality reduction analysis (Fig. 4a, b) clearly delineated tumour and normal regions, with HU areas concentrated at the boundaries in feature space. We were able to effectively extract these areas through uncertainty quantification.

**Figure 4.**
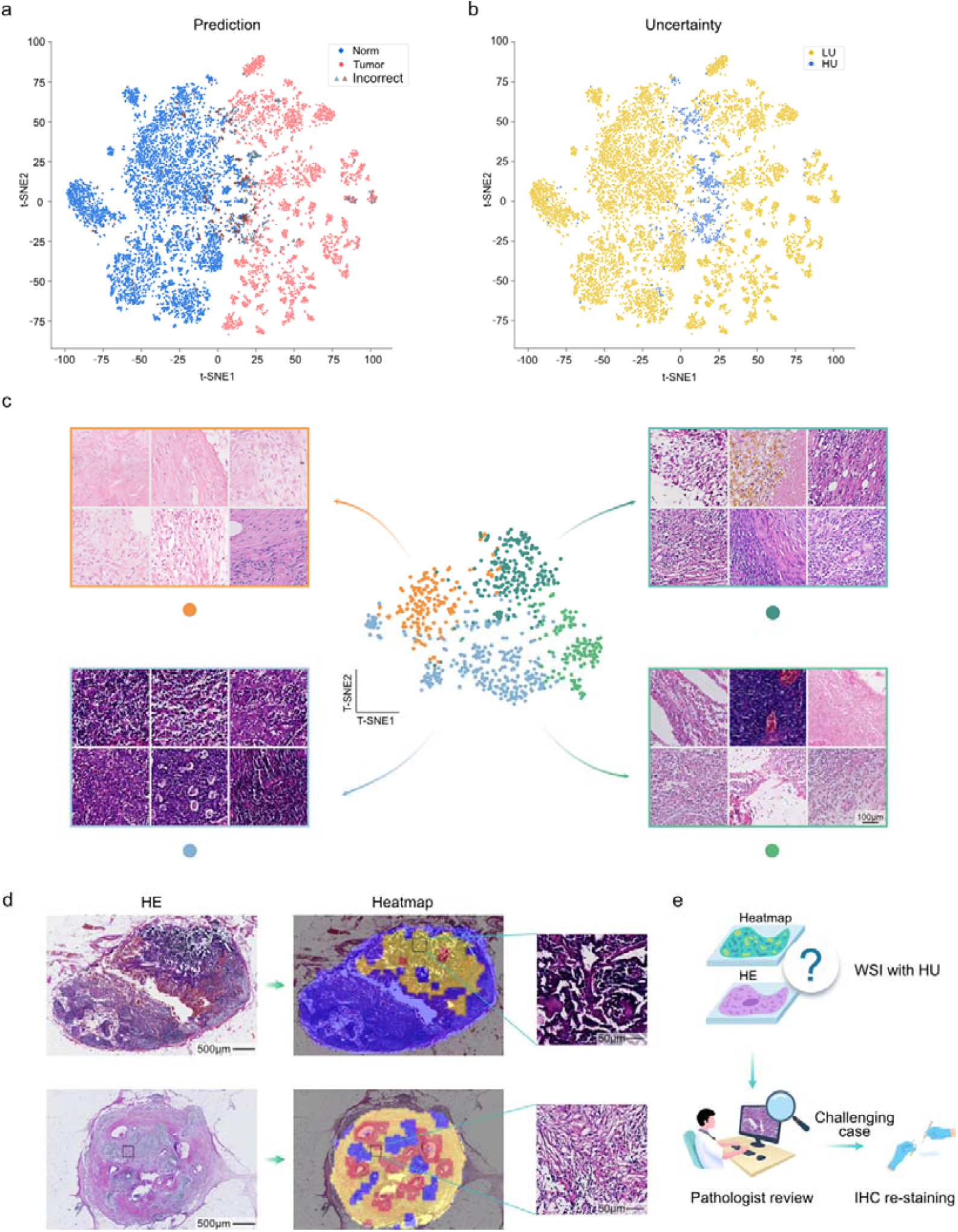
Visualization of uncertainty estimation. (a-b) t-SNE visualization of model features from internal validation dataset: (a) Tiles colored by binary tumor prediction (tumor/normal) with triangles indicating misclassified samples (incorrect); (b) Tiles colored by uncertainty levels (blue: low (LU), yellow: high (HU)). (c) Cluster analysis of HU tiles showing representative tiles from four categories: dense connective tissue, loose connective tissue, low-quality image, and bleeding. (d) Representative HU LN cases highlighting HU regions (yellow) and tumor regions (red). (e) Expert validation of HU predictions with IHC confirmation for challenging cases.

Further analysis allowed categorization of HU regions into types such as dense connective tissue, loose connective tissue, low-quality image areas, and bleeding areas (Fig. 4c). As shown in Fig. 4d, the model assigned high HU scores to two patients with distinct atypical cytomorphology: one with lymphocyte-like tumour cells featuring scant cytoplasm and naked nuclei, and another exhibiting plasmacytoid/histiocytoid features with moderate cytoplasm. This assessment mechanism enables pathologists to selectively focus on these challenging cases, thereby ensuring diagnostic accuracy and clinical safety while improving efficiency. As the system continues to be applied, manual review of uncertain regions will facilitate ongoing model optimization (Fig. 4e).

## Discussion

This study addresses the limitations of current organ-specific AI paradigms by presenting UPATHLN, a unified, uncertainty-aware framework for pan-cancer LNM diagnosis. While previous deep learning approaches have validated LNM detection in organ-specific cohorts, they often operate as “black boxes,” lacking the safety mechanisms required for broad clinical adoption^21^. By integrating a pathology foundation model with decoupled uncertainty estimation, UPATHLN establishes a robust clinical safety net. Crucially, the system demonstrated a “fail-safe” capability in a large-scale independent cohort, where the uncertainty mechanism successfully intercepted all missed metastases to achieve 100% conditional sensitivity. This achievement marks a significant advancement: moving from algorithms that prioritize raw accuracy to safety-critical systems that effectively mitigate the risk of false negatives in metastatic screening.

The uncertainty bypass architecture serves as the technical foundation of this safety profile. Classical uncertainty estimation methods—such as uncertainty-prediction model co-training^29,30^, multiple model ensembles^31^, or repeated dropout strategies^29,32^—increase computational burden and often flag large unnecessary regions for manual reassessment. By contrast, our independent bypass architecture provides precise localization of diagnostically uncertain regions without compromising inference speed. This decoupled mechanism acts as a reliable diagnostic filter, specifically designed to handle the inherent biological heterogeneity of metastatic disease. It respects the ambiguity of “long-tail” rare variants and non-neoplastic confounders (e.g., anthracotic pigment), routing these cases strictly within the system’s competence boundaries. This “rescue effect” ensures that algorithmic limitations do not translate into diagnostic errors, maintaining high reliability even when encountering complex morphologies that typically confound conventional models.

Furthermore, we successfully navigated the data annotation bottleneck that typically constrains the scalability of foundation models. By coupling high-level feature representations with an uncertainty-guided active learning framework, we efficiently captured a comprehensive spectrum of metastatic morphologies across 14 cancer types without necessitating exhaustive annotation. This strategy proved vital for addressing the diverse patterns of metastatic pathology. Consequently, the system maintained robust performance even on LNM samples from primary sites entirely unseen during training. This demonstrates that uncertainty-driven learning provides a practical, scalable pathway to generalize pathology AI, enabling adaptability to domain shifts and rare cancer types lacking large-scale training data.

The translational implication of UPATHLN lies in its ability to optimize clinical workflows without compromising safety. Our analysis indicates that the system can safely automate the exclusion of 73.2% of negative LNs. This establishes a highly efficient synergistic workflow: the AI handles the bulk screening of negative slides, allowing pathologists to focus their expertise on the remaining subset of positive or high-uncertainty cases. Moreover, by enabling the precise quantification of metastatic burden beyond crude pN staging, UPATHLN provides a basis for the refined quantitative assessment required for modern precision oncology^13^.

Despite these advances, limitations remain. First, as a retrospective study, the operational integration of this workflow warrants validation in prospective multi-centre trials. Second, while artifacts are effectively flagged as uncertain, future iterations could incorporate specific suppression modules to further reduce the review burden. In conclusion, UPATHLN establishes a robust, scalable, and safe framework for pan-cancer LNM diagnostics. By harmonizing automation efficiency with a built-in safety net, it overcomes the fragmentation of existing tools and sets a new standard for trustworthy, unified AI in clinical practice.

## Data availability

The data of CAMELYON16 (BLN16) is publicly available via the Grand Challenge platform (https://camelyon16.grand-challenge.org). The remaining datasets are not publicly available due to hospital regulations and patient privacy. Requests for access to these restricted raw data should be directed to the corresponding author.

## Code availability

The lymph node detection was conducted using the YOLOv8-s model (https://github.com/ultralytics/ultralytics), while segmentation utilized nnUNet (https://github.com/MIC-DKFZ/nnUNet). The pathology foundation model employed was UNI (https://github.com/mahmoodlab/UNI). The classification model will be publicly available on GitHub upon acceptance. For more details on additional tools and libraries, please refer to the Methods section.

## Ethics oversight

This study was approved by the Ethics Committee of Changhai Hospital (CHEC2024-276), the Ethics Committee of the Second People’s Hospital of Wuxi (2024Y-125), and the Ethics Committee of the 960th Hospital of the PLA (2024-087). The requirement for informed consent was waived due to the retrospective nature of the study and the use of anonymized data.

## References

1. Siegel, R. L., Kratzer, T. B., Giaquinto, A. N., Sung, H. & Jemal, A. Cancer statistics, 2025. CA. Cancer J. Clin. 75, 10–45 (2025).

2. Mani, K. et al. Causes of death among people living with metastatic cancer. Nat. Commun.15, (2024).

3. Dillekås, H., Rogers, M. S. & Straume, O. Are 90% of deaths from cancer caused by metastases? Cancer Med. 8, 5574–5576 (2019).

4. Ji, H. et al. Lymph node metastasis in cancer progression: molecular mechanisms, clinical significance and therapeutic interventions. Signal Transduct. Target. Ther. 8, 1–33 (2023).

5. Reticker-Flynn, N. E. et al. Lymph node colonization induces tumor-immune tolerance to promote distant metastasis. Cell 185, 1924-1942.e23 (2022).

6. TNM Classification of Malignant Tumours. (John Wiley & Sons, Inc, Chichester, West Sussex, UK Hoboken, NJ, 2017).

7. Aoyama, T. et al. Clinical influence of the lymph node ratio on lymph node metastasis-positive gastric cancer patients who receive curative treatment. Vivo Athens Greece 36, 994–1000 (2022).

8. Jones, D., Pereira, E. R. & Padera, T. P. Growth and immune evasion of lymph node metastasis. Front. Oncol. 8, 36 (2018).

9. Herrada, A. A. et al. Lymph Leakage Promotes Immunosuppression by Enhancing Anti-Inflammatory Macrophage Polarization. Front. Immunol. 13, 841641 (2022).

10. Bray, F. et al. Global cancer statistics 2022: GLOBOCAN estimates of incidence and mortality worldwide for 36 cancers in 185 countries. CA. Cancer J. Clin. 74, 229–263 (2024).

11. Wu, S. et al. Artificial intelligence-based model for lymph node metastases detection on whole slide images in bladder cancer: a retrospective, multicentre, diagnostic study. Lancet Oncol. 24, 360–370 (2023).

12. Ehteshami Bejnordi, B. et al. Diagnostic assessment of deep learning algorithms for detection of lymph node metastases in women with breast cancer. JAMA 318, 2199 (2017).

13. Wang, X. et al. Predicting gastric cancer outcome from resected lymph node histopathology images using deep learning. Nat. Commun. 12, 1637 (2021).

14. Pan, Y. et al. Automatic detection of squamous cell carcinoma metastasis in esophageal lymph nodes using semantic segmentation. Clin. Transl. Med. 10, e129 (2020).

15. Pham, H. H. N. et al. Detection of lung cancer lymph node metastases from whole-slide histopathologic images using a two-step deep learning approach. Am. J. Pathol. 189, 2428–2439 (2019).

16. Song, J. H., Hong, Y., Kim, E. R., Kim, S.-H. & Sohn, I. Utility of artificial intelligence with deep learning of hematoxylin and eosin-stained whole slide images to predict lymph node metastasis in T1 colorectal cancer using endoscopically resected specimens; prediction of lymph node metastasis in T1 colorectal cancer. J. Gastroenterol. 57, 654–666 (2022).

17. Lenharo, M. An AI revolution is brewing in medicine. What will it look like? Nature 622, 686–688 (2023).

18. Moor, M. et al. Foundation models for generalist medical artificial intelligence. Nature 616, 259–265 (2023).

19. Guha Roy, A. et al. Does your dermatology classifier know what it doesn’t know? Detecting the long-tail of unseen conditions. Med. Image Anal. 75, 102274 (2022).

20. Fujii, M., Sekine, S. & Sato, T. Decoding the basis of histological variation in human cancer. Nat. Rev. Cancer 24, 141–158 (2024).

21. Fleisher, L. A. & Economou-Zavlanos, N. J. Artificial intelligence can Be regulated using current patient safety procedures and infrastructure in hospitals. JAMA Health Forum 5, e241369 (2024).

22. Ellahham, S., Ellahham, N. & Simsekler, M. C. E. Application of artificial intelligence in the health care safety context: opportunities and challenges. Am. J. Med. Qual. 35, 341–348 (2020).

23. Xiang, J. et al. A vision–language foundation model for precision oncology. Nature 1–10 (2025) doi:10.1038/s41586-024-08378-w.

24. Wang, X. et al. A pathology foundation model for cancer diagnosis and prognosis prediction. Nature 1–9 (2024) doi:10.1038/s41586-024-07894-z.

25. Chen, R. J. et al. Towards a general-purpose foundation model for computational pathology. Nat. Med. 30, 850–862 (2024).

26. Xu, H. et al. A whole-slide foundation model for digital pathology from real-world data. Nature 630, 181–188 (2024).

27. Lu, M. Y. et al. A visual-language foundation model for computational pathology. Nat. Med. 30, 863–874 (2024).

28. Vorontsov, E. et al. A foundation model for clinical-grade computational pathology and rare cancers detection. Nat. Med. 30, 2924–2935 (2024).

29. Isensee, F., Jaeger, P. F., Kohl, S. A. A., Petersen, J. & Maier-Hein, K. H. nnU-net: a self-configuring method for deep learning-based biomedical image segmentation. Nat. Methods 18, 203–211 (2021).

30. Dosovitskiy, A. et al. An image is worth 16x16 words: transformers for image recognition at scale. Preprint at 10.48550/ARXIV.2010.11929 (2020).

31. Pan, Y. et al. Clinically applicable pan-origin cancer detection for lymph nodes via artificial intelligence-based pathology. Pathobiology 1–14 (2024) doi:10.1159/000539010.

32. Linmans, J., Elfwing, S., Van Der Laak, J. & Litjens, G. Predictive uncertainty estimation for out-of-distribution detection in digital pathology. Med. Image Anal. 83, 102655 (2023).

33. Linmans, J., Hoogeboom, E., Van Der Laak, J. & Litjens, G. The latent doctor model for modeling inter-observer variability. IEEE J. Biomed. Health Inform. 1–12 (2023) doi:10.1109/JBHI.2023.3323582.

34. Olsson, H. et al. Estimating diagnostic uncertainty in artificial intelligence assisted pathology using conformal prediction. Nat. Commun. 13, 7761 (2022).

35. Dolezal, J. M. et al. Uncertainty-informed deep learning models enable high-confidence predictions for digital histopathology. Nat. Commun. 13, 6572 (2022).

